# Real-World Performance of Urine β-amyloid Test Kits in Multiple Hospital Clinics and Neighborhood Communities of China

**DOI:** 10.64898/2026.05.06.26348372

**Authors:** Liyan Qiao, Guihong Wang, Xiaotong Chen, Jing Wang, Wei Huang, Dongmei Xing, Qingfang Zhao, Yunliang Wang, Honglei Yin, Houozhen Tuo, Shiya Wang, Guanghong Xiang, Nina Zhou, Yong Lin, Jun Wang, Hongzheng Wang

## Abstract

**Background:** Growing evidence suggests that urinary β-amyloid precursor protein (AβPP) fragments can serve as an early screening biomarker for mild cognitive impairment and dementia. However, in reality, older adults, regardless of the presence of cognitive decline, often suffer from multiple age-related conditions and are on multiple medications. How these comorbidities and treatments affect the performance of early diagnostic biomarkers remains unclear.

**Methods:** This study further validated the sensitivity, specificity, and clinical value of the Qankorey® urinary β-amyloid protein detection kit in early dementia screening through a randomized community screening (n=51187) conducted in Changsha, and a multicenter case-control study conducted at Yuquan Hospital (Tsinghua University), Tiantan Hospital (Capital Medical University), Beijing Friendship Hospital, Zibo 148 Hospital (Shandong), and the Third People’s Hospital of Yunnan Province. The multicenter case-control study included 898 participants, comprising 266 healthy, age-matched controls without any comorbidities, 167 patients with mild cognitive impairment/Alzheimer’s disease (MCI/AD), and 465 non-AD patients with various comorbidities and age-related diseases.

**Results:** The kit showed a significant age-dependent positive rate in both men and women in Changsha, increasing from 6.29% to 15.40%. The number of weakly positive/positive/negative individuals in the healthy group, non-AD group, and MCI/AD group were 8/12/246 (positive rate 7.52%), 41/16/409 (12.23%), and 77/44/46 (72.46%), respectively, with a Kappa value of 0.669, indicating that the method performed well in the clinical diagnosis of MCI/AD, consistent with previously published results. Among the 8 weakly positive healthy subjects, 6 were found to have brain abnormalities by MRI/CT examination. Comorbidity analysis showed that memory decline was the most significantly associated risk factor (P=9.6×10 □^23^, Fisher’s exact test), followed by hyperlipidemia (P=3.2×10 □^12^), history of stroke (P=0.0011), and hypertension (P=0.0058). Treatment analysis showed that cardiovascular drugs and antithrombotic drugs significantly reduced the risk association with dementia (P values were 0.0061 and 0.0081, respectively), followed by hypoglycemic drugs (P=0.0358). For AD patients, those receiving only memantine showed a slightly lower positive test rate (P=0.0532).

**Conclusion:** Our findings confirm the diagnostic value of urinary β-amyloid protein detection in MCI and AD-related dementia. Furthermore, this kit can be used in practical clinical applications to assess the risk of cognitive decline and treatment efficacy across various diseases.

## Introduction

Alzheimer’s disease (AD) is becoming a greater burden on the global aging population. It is estimated that by 2050, there will be 114 million to 150 million people with AD worldwide ^[1]^. There is currently no effective cure for AD. Existing treatments are all disease-modifying therapies, which are only suitable for early-stage patients and can only slow the progression of the disease. Once medication is stopped, the symptoms will worsen. This makes early prevention and early intervention the key to controlling the onset of AD.

The incubation period of AD can be as long as 20-30 years ^[2]^. Identifying high-risk groups during the incubation period, before the onset of clinical symptoms, has remained an unsolved problem. Traditional AD diagnosis relies on cognitive tests and imaging examinations, such as Aβ-PET scans and magnetic resonance imaging (MRI). Cognitive tests are too subjective and are greatly affected by the level of education. PET scans are expensive and require the injection of radioactive traces into the body, making it unsuitable for large-scale screening. Recently emerging blood biomarkers such as the Aβ 40/42 ratio, p-Tau181, and p-Tau217 have provided the possibility of early screening. Changes in Tau are generally recognized by researchers as secondary changes in Aβ ^[2,3]^, and therefore are not suitable as an early screening biomarker. In addition, the detection of these biomarkers generally requires expensive instruments and reagents, and is not suitable for large-scale primary screening. However, they could play a greater role in further examination of positive individuals identified in primary screening.

β-amyloid precursor protein (AβPP) is a protein that protects neural synapses ^[4,5]^ and regulates neuronal excitability, synaptic activity, signal transmission and plasticity. Its secretion increases when neurons are stressed or damaged ^[6-8]^. Increased AβPP is found not only in the cerebrospinal fluid (CSF) ^[9]^ and blood ^[10]^ of AD patients (especially those who do not carry APOE ε4), but also in other types of neurodegenerative diseases such as motor neuron disease ^[11,12]^. Therefore, elevated AβPP is a biomarker of impaired brain health, not just AD. Persistently elevated AβPP may indicate persistent brain problems and could potentially lead to AD or other types of dementia. Aβ plaques form in the brain of AD patients due to disturbed innate phagocytic function which creates difficulty in clearance of AβPP fragments ^[13]^. Other types of dementia patients do not form plaques if their innate phagocytic function is undisturbed, but this does not prevent the development of other pathologies. One of the main pathological features of vascular dementia (VaD) is the deposition of Aβ plaques in blood vessel wall layers ^[14]^. Patients with frontotemporal dementia (FTD) and dementia with Lewy body (DLB) also exhibit similar pathological features to AD as their innate phagocytic capacity decreases with aging, such as Aβ plaques ^[15,16]^. Therefore, elevated AβPP, as a normal response of living nerve cells, can appear at the source of almost all types of dementia pathology. Other biomarkers, such as neurofilament light chain (NfL), Alzheimer-associated neuronal thread protein (AD7c-NTP), and Tau are leaked following nerve cell death, and their early screening significance is far less than that of AβPP fragments. This is because large-scale nerve cell death is unlikely to occur in the early stages of disease.

As one of the degradation products of AβPP, Aβ is the earliest known biomarker of AD. It begins to accumulate 20-30 years before the onset of clinical symptoms. In humans, this manifests as an increase in the metabolism of Aβ and its precursor protein AβPP, including the formation of Aβ plaques in brain ^[2,3]^ and increase of AβPP fragments in blood and urine ^[17,18]^, supporting its potential to be used as an early screening biomarker. However, because the content of Aβ in urine is extremely low and it quickly forms oligomers, and AβPP in the body is easily degraded by enzymes, it is very difficult to accurately measure their concentration in urine ^[19]^. Qankorey Biotechnology has launched the world’s first urine β-amyloid protein detection kit ^[20]^. It uses a unique antibody-antigen-polymer high-sensitivity sandwich method for detection. Published data shows that it can distinguish mild cognitive impairment (MCI) patients who have not yet developed Aβ plaques. Moreover, it performed well in the screening of 11 cities in Jiangsu Province organized by the Jiangsu Provincial Health Commission in 2023 ^[20]^. Since there is strong demand for a simple and inexpensive biomarker-based tool that can be used for primary community screening, it is necessary to determine whether this product meets those needs. Meanwhile, the product can also be used for self-testing at home, which can supplement the shortcomings of community screening and protect personal privacy ^[21]^.

Besides the lack of suitable screening methods, another challenge in screening for cognitive decline risk in the elderly is the impact of comorbidities and medications on screening results. In reality, most older adults have various common illnesses, such as high blood sugar, high blood pressure, high cholesterol, and memory loss. It is unknown what confounding impacts, if any, these common illnesses and their medications have on cognitive decline or improvement. Currently, there are no good methods to measure and assess these factors. In this study, we collected relevant information to try and determine any impact of those factors.

## Methods

### Multicenter validation

Yuquan Hospital of Tsinghua University, Beijing Tiantan Hospital and Beijing Friendship Hospital of Capital Medical University, Zibo No. 148 Hospital in Shandong Province, and the Third People’s Hospital of Yunnan Province participated in this work. Participants were over 55 years of age, regardless of gender. The healthy control group consisted of participants without any known age-related diseases and who did not regularly take medications. The mild cognitive impairment (MCI) and Alzheimer’s disease (AD) groups consisted of clinically diagnosed outpatients or inpatients with MCI or AD, the vast majority of whom were undergoing treatment. The non-AD disease group consisted of elderly individuals with diseases (excluding MCI or AD) but normal cognition, the vast majority of whom were continuously taking medications. The Mini-Mental State Examination (MMSE), the Montreal Cognitive Assessment (MoCA), and the Qiankorey® Urine β-Amyloid Protein Detection Kit (also known as “One-Step Dementia Risk Test Kit”) developed by Qankorey Biotechnology, Changsha, Hunan Province, China, were used for testing. Some participants underwent multiple tests with varying intervals between tests, approximately 7-30 days. In cases of inconsistency among the three results, the majority result was considered valid. This study was approved by the Ethics Committee of Yuquan Hospital of Tsinghua University (Tsinghua University Hospital of Integrated Traditional and Western Medicine) (Approval No. 2023015).

### Community Screening

In response to the national call to promote the prevention and control of AD, the Yuhua District People’s Government and the Yuhua Economic Development Zone Management Committee of Changsha City, Hunan Province, actively coordinated and organized the screening program. The Yuhua District Health and Wellness Bureau and the Yuhua District Center for Disease Control and Prevention provided on-site coordination, meticulous organization, and full mobilization. Expert support was provided by the Second People’s Hospital of Hunan Province (Hunan Provincial Brain Hospital), and early screening kits for dementia were provided by Qankorey Biotechnology. From March to December 2024, free early screening and prevention services for dementia were conducted for individuals aged 50 and above in 14 community health service centers/hospitals within the Yuhua district. A total of 51,187 people were tested.

### Statistical methods

We used the chi-square test (χ^2^ test ) and Fisher’s exact test to detect differences between groups. The Kappa test was used to evaluate the clinical diagnostic value of the test strips: 0.81–1 was highly consistent, 0.61–0.8 was good consistent, 0.41–0.6 was moderately consistent, 0.21–0.4 was reasonably consistent, and < 0.2 was poor.

## Results

### 1. Summary Data from Multicenter Clinical Trials

A total of 898 participants were enrolled in the multicenter clinical trials, mostly aged 60-75. This included 266 healthy, age-matched controls with no known age-related diseases, 167 patients with MCI or AD, and 465 patients with various diseases and poor health (Table 1). These diseases primarily included: hypertension, hyperglycemia, hyperlipidemia, history of stroke, heart disease, Parkinson’s disease, sleep disorders, dizziness, memory loss, headaches and other brain disorders, partial limb weakness and numbness, inflammation and infection, and anxiety. Nearly half of the participants (n = 415) underwent three tests, with 391 showing consistent initial and final results (94.2% concordance rate), indicating good stability of the test strip results. The positive rates for the healthy control group, MCI/AD patient group, and age-related disease group were 7.52%, 72.46%, and 12.23%, respectively (Table 1). Comparing the results of the healthy control (HC) group and the MCI/AD patient group, the positive concordance rate (sensitivity) of the test strips was 72.5%, the negative concordance rate (specificity) was 92.5%, the theoretical concordance rate was 54%, and the actual concordance rate was 84.8%, with a Kappa (κ) value of 0.669 (Table 1), showing a good clinical concordance and diagnostic value, consistent with previous reports ^[20]^. While evaluating its early screening value, we compared the HC group with the MCI group: the sensitivity was 79.2%, specificity was 92.5%, the theoretical concordance rate was 80.5% and the actual concordance rate was 91.4%, Kappa (κ) value was 0.558, showing its good potential for early screening. All 8 participants in the healthy control group with weak positive test strips were arranged to undergo further MRI and CT examinations, and 6 of them were found to have brain abnormalities (see supplementary material “Mastersheet_En.xlsx”).

**Table 1:** Summary Data of Multicenter Clinical Trials.

|  | HC | Non-AD | MCI | AD | MCI+AD |
| --- | --- | --- | --- | --- | --- |
| Number (male/female) | 266 (107/159) | 465(237/228) | 24 ( 12/12 ) | 143 ( 50/93 ) | 167(62/105) |
| Age ($\bar{x} \pm s$) | 65.4±10.3 | 67.0±8.4 | 70.7±8.2 | 70.7±9.8 | 70.7±9.6 |
| Weak positive | 8 | 41 | 16 | 61 | 77 |
| Positive | 12 | 16 | 3 | 41 | 44 |
| Negative | 246 | 409 | 5 | 41 | 46 |
| % Positive | 7.5% | 12.2% | 79.2% | 71.3% | 72.5% |
| P value | - | 0.0286 | $2.60 \times 10^{-13}$ | $1.01 \times 10^{-41}$ | $1.14 \times 10^{-46}$ |

Note: P values were calculated using Fisher's exact test compared with the healthy control (HC) group.

### 2. Test strip results are closely correlated to cognition

We then examined the performance of the test strips in different cognitive populations. Based on MMSE and MoCA scores, we divided the subjects into four groups: normal cognitive group (MMSE >26, MoCA >25), mild cognitive decline group (MMSE 23-26, MoCA 18-25), moderate cognitive decline group (MMSE 10-22, MoCA 10-17) and severe cognitive decline group (MMSE <10, MoCA <10) (Table 2). The results showed that the positive rate of the test strips increased significantly with decreasing MMSE and MoCA scores, indicating that they can accurately reflect cognitive decline.

**Table 2:** Performance of urine Aβ test strips in different cognitive populations.

|  | MMSE score |  |  |  |
| --- | --- | --- | --- | --- |
|  | 27-30 | 23-26 | 10-22 | <10 |
| Test strip positive | 69 | 18 | 67 | 43 |
| Test strip negative | 515 | 47 | 115 | 15 |
| Positive rate | 11.8% | 27.7% | 36.8% | 74.1% |
| p-value | 0.0009 | 0.1190 | 5.76$\times 10^{-11}$ | |

|  | MoCA score |  |  |  |
| --- | --- | --- | --- | --- |
|  | >25 | 18-25 | 10-17 | <10 |
| Test strip positive | 35 | 50 | 25 | 34 |
| Test strip negative | 376 | 194 | 52 | 17 |
| Positive rate | 8.5% | 20.5% | 32.5% | 66.7% |
| p-value | 1.23$\times 10^{-11}$ | 0.0248 | 0.0001 | |

Note: P-values are Fisher's Exact Test results compared to the adjacent right group.

### 3. The impact of ApoE genotyping

ApoE ε4 genotyping is a known high-risk factor for AD. We examined the positivity rates of test strips in ε4^+^ (ε2/ε4, ε3/ε4 and ε4/ε4) and ε4^-^ (ε2/ε2, ε2/ε3 and ε3/ε3) subjects and found no significant difference in positivity rates between ε4 carriers and non-carriers in cognitively normal individuals (healthy controls and non-AD groups) and cognitively impaired individuals (MCI and AD groups) (Table 3). This may be due to insufficient sample size. Notably, in the cognitively normal population, all 9 ε4 carriers tested negative. Previous literature has shown that healthy individuals carrying ε4 have a lower positivity rate than those without ε4, possibly related to the fact that ApoE4 protein binds more readily to Aβ ^[22,23]^. Our results, to some extent, validated the previous report.

**Table 3:** Urine Aβ test strips in ApoE Performance in ε4 and non-ε4 populations.

|  | Cognitively Normal |  | MCI+AD |  | Everyone |  |
| --- | --- | --- | --- | --- | --- | --- |
| | $\epsilon 4^+$ | $\epsilon 4^-$ | $\epsilon 4^+$ | $\epsilon 4^-$ | $\epsilon 4^+$ | $\epsilon 4^-$ |
| Test strip positive | 0 | 6 | 25 | 26 | 25 | 32 |
| Test strip negative | 9 | 26 | 19 | 13 | 28 | 39 |
| Positive rate | 0 | 18.7% | 56.8% | 66.7% | 47.2% | 45.1% |
| p-value | 0.2015 |  | 0.2441 |  | 0.4798 |  |

### 4. The effect of a single comorbidity on test strips

We then analyzed the impact of different comorbidities on the positive rate of the test strips by comparing individuals with and without comorbidities (Table 4). These AD-related comorbidities and sub-health conditions included hypertension, hyperglycemia, hyperlipidemia, history of stroke, heart disease, Parkinson’s disease, sleep disorders, dizziness, memory loss, headaches and other brain diseases, limb weakness, inflammatory infections, and anxiety. The results showed that, if the risk of dementia was measured by the positive rate of the test strips, individuals with memory loss were the most at risk (P = 9.67 × 10^-23^, Fisher’s Exact Test, same below), followed by hyperlipidemia (P = 3.25 × 10 □^12^), history of stroke (P = 0.0011) and hypertension (P = 0.0058). Surprisingly, hyperglycemia (P = 0.1739) is not included in this category, which contradicts the common understanding that AD could be a type III diabetes. However, this result may explain the failure of the clinical trials (EVOKE and EVOKE PLUS) targeting blood sugar reduction for AD announced in November 2025. To our surprise, people with frequent dizziness apparently showed lower positivity rate of the test strip (P = 1.26 × 10 □ ^1^ □), probably due to the associated hypotension.

**Table 4-1:** Performance of urine Aβ test strips in all populations with or without different comorbidities.

| Comorbidities | comorbidities |  |  | No comorbidities |  |  | P |
| --- | --- | --- | --- | --- | --- | --- | --- |
|  | Pos | Neg | Positive rate | Pos | Neg | Positive rate |  |
| Hypertension | 106 | 302 | 26.0 % | 92 | 399 | 18.7 % | **0.0058** |
| Hyperglycemia | 37 | 155 | 19.3 % | 161 | 546 | 22.8 % | 0.1739 |
| Hyperlipidemia | 73 | 96 | 43.2 % | 125 | 605 | 17.1 % | **$3.25 \times 10^{-12}$** |
| History of Stroke | 22 | 32 | 40.7 % | 176 | 669 | 20.8 % | **0.0011** |
| Heart disease | 11 | 56 | 16.4 % | 187 | 645 | 22.5 % | 0.1590 |
| Parkinson's disease | 3 | 8 | 27.3 % | 195 | 693 | 22.0 % | 0.4499 |
| Sleep disorders | 6 | 20 | 23.1 % | 192 | 681 | 22.0 % | 0.5260 |
| Frequent dizziness | 38 | 343 | 10.0 % | 159 | 358 | 30.8 % | **$1.26 \times 10^{-14}$** |
| Memory loss | 102 | 112 | 47.7 % | 96 | 589 | 14.0 % | **$9.67 \times 10^{-23}$** |
| Brain diseases | 5 | 13 | 27.8 % | 193 | 688 | 21.9 % | 0.3617 |
| Limb weakness | 22 | 94 | 19.0 % | 176 | 607 | 22.5 % | 0.2346 |
| Inflammation | 2 | 15 | 11.8 % | 196 | 686 | 22.2 % | 0.2399 |
| Anxiety | 6 | 6 | 50.0 % | 192 | 695 | 21.6 % | **0.0297** |

If we only analyze cognitively normal individuals, including healthy individuals and those without AD disease, old cerebral infarction and hyperlipidemia are the most associated risk factors, with P values of 7.39 × 10□□ and 6.34 × 10□□, respectively. This is followed by hypertension (P = 0.0027), anxiety (P = 0.0095), and sleep disorders (P = 0.0461) (Table 4-2). Memory decline was not significantly associated with cognitive decline/AD in cognitively normal individuals, possibly because this symptom is too subjective, and clinical descriptions do not distinguish between general memory decline and episodic memory decline. Therefore, it is difficult to effectively differentiate between age-related normal memory decline and pathological memory decline in early-stage dementia. We also found elderly individuals with frequent dizziness in cognitively normal individuals, and the positive rate of the test kit was significantly lower than that of elderly individuals without this symptom (P = 0.0028).

**Table 4-2:** Performance of urine Aβ test kit in cognitively normal participants with or without different comorbidity comorbidities No comorbidities.

| Comorbidities | comorbidities |  |  | No comorbidities |  |  | P |
| --- | --- | --- | --- | --- | --- | --- | --- |
|  | Pos | Neg | Positive rate | Pos | Neg | Positive rate |  |
| Hypertension | 46 | 277 | 14.2 | 31 | 378 | 7.6 | 0.0027 |
| Hyperglycemia | 19 | 148 | 11.4 | 58 | 507 | 10.3 | 0.3871 |
| Hyperlipidemia | 23 | 77 | 23.0 | 54 | 578 | 8.5 | $6.34 \times 10^{-5}$ |
| History of Stroke | 16 | 32 | 33.3 | 61 | 623 | 8.9 | $7.39 \times 10^{-6}$ |
| Heart disease | 7 | 55 | 11.3 | 70 | 600 | 10.4 | 0.4845 |
| Parkinson's disease | 2 | 7 | 22.2 | 75 | 648 | 10.4 | 0.2427 |
| Sleep disorders | 6 | 20 | 23.1 | 71 | 735 | 10.1 | 0.0461 |
| Frequent dizziness | 27 | 343 | 7.3 | 50 | 312 | 13.8 | 0.0028 |
| Memory loss | 102 | 112 | 13.9 | 96 | 589 | 10.1 | 0.1942 |
| Brain diseases | 5 | 13 | 18.8 | 193 | 688 | 10.3 | 0.2319 |
| Limb weakness | 22 | 94 | 8.8 | 176 | 607 | 10.8 | 0.3447 |
| Inflammation | 2 | 15 | 11.8 | 196 | 686 | 10.5 | 0.5493 |
| Anxiety | 6 | 6 | 44.4 | 192 | 695 | 10.1 | 0.0095 |

**Table 5:** Performance of urine Aβ test strips in populations taking different medications.

| Treatment medication | Strip Results |  |  | P |
| --- | --- | --- | --- | --- |
|  | Pos | Neg | Pos rate |  |
| **Non-AD treatment drugs** | 17 | 359 | 4.5% | 4.53 ×<br>10<sup>-4</sup> |
| Antithrombotic | 4 | 96 | 4.0% | 0.0081 |
| Lipid-lowering | 9 | 102 | 8.1% | 0.1433 |
| Antihypertensive | 11 | 140 | 7.3% | 0.0581 |
| Hypoglycemic agents | 2 | 53 | 3.6% | 0.0358 |
| Brain-improving | 2 | 31 | 6.1% | 0.2249 |
| Cardiovascular | 0 | 41 | 0 | 0.0060 |
| Dizziness and tinnitus | 0 | 36 | 0 | 0.0110 |
| **AD treatment drugs** | 72 | 38 | 65.5% | 0.1303 |
| Memantine hydrochloride | 25 | 18 | 58.1% | 0.0532 |
| GV-971 | 65 | 34 | 65.7% | 0.1514 |
| Donepezil hydrochloride | 41 | 12 | 77.4% | 0.3031 |

Note: P-values are Fisher's Exact Test results. Non-AD treatment drugs are compared with the non-AD disease group in Table 1 (positive number 57, negative number 409), and AD treatment drugs are compared with the MCI+AD group in Table 1 (positive number 121, negative number 46).

### 5. Effects of Medications

We analyzed medications commonly used by the elderly. Medications are categorized into **antithrombotic drugs** (aspirin, clopidogrel, indobufen, alteplase, ticagrelor, argatroban and rivaroxaban), **lipid-lowering drugs** (atorvastatin, rosuvastatin, pitavastatin, simvastatin, acilimus and probucol), **antihypertensive drugs** (amlodipine, losartan potassium, nifedipine, amlodipine, valsartan hydrochlorothiazide, enalapril, irbesartan, bisoprolol fumarate, felodipine, carvedilol, captopril), **hypoglycemic drugs** (metformin, acarbose, glimepiride, dapagliflozin, insulin), **neurostimulants** (huperzine A tablets, nicergoline, blood-nourishing and brain-clearing drugs, vitamin B1, methylcobalamin, nerve growth factor, citicoline sodium tablets, idebenone, piribedil, folic acid tablets), and **antidepressants** (escitalopram oxalate tablets, duloxetine (flupentixol melitracen), mirtazapine tablets, risperidone, duloxetine hydrochloride), **cardiovascular/stroke drugs** (ginkgo diterpenes, nicotinic acid injection, danshen injection, sanqitongshu, tanshinone polyphenolic acid, edaravone dexborneol, butylphthalide, ginkgolide injection, sacubitril valsartan sodium tablets), and **dizziness and tinnitus drugs** (betahistine). In the medication analysis of non-AD patients, the positive rate of test strips in the group that regularly took medications was lower than that in the non-AD disease group, and in some cases even lower than that in the normal control group. Cardiovascular disease drugs and antithrombotic drugs significantly reduced the risk of cognitive decline (P values were 0.006 and 0.008, respectively), followed by hypoglycemic drugs (P = 0.036). In the medication analysis of drugs for AD, the positive rate of test strips in patients receiving memantine hydrochloride was slightly lower than the overall positive rate of the patients (P = 0.0558), while there was no significant difference between commonly used GV-971 and donepezil hydrochloride tablets.

### 6. Community screening

The subsequent large-scale community screening received strong support from the Health Commission of Yuhua District and the leaders of the Economic Development Zone in Changsha City. A total of 51,187 middle-aged and elderly people participated in the screening. The positive rate of the test strips increased with different age groups, ranging from 7.8% (46-59 years old) to 15.4% (over 89 years old) (Table 6), which is consistent with the incidence trend of AD in different age groups in China. For the 4,900 participants who tested positive, each health service center/hospital notified them to undergo re-testing, but only 449 participants actually did. Among them, five participants who tested positive twice voluntarily participated in subsequent hospital examinations, and all of them were found to have brain atrophy or were diagnosed with early AD. This pilot screening project shows that the urinary β-amyloid protein detection kit, as a novel, non-invasive, rapid, and low-cost tool for early screening and auxiliary diagnosis of AD, has great feasibility in the real world.

**Table 6:** Results of Community-wide Screening in Yuhua District.

| Age group | Number of people (male/female) | Number of negative | Number of positive | Positive rate (%) |
| --- | --- | --- | --- | --- |
| < 46 | 334 (188/146) | 313 (176/137) | 21 (12/9) | 6.3 (6.4/6.2) |
| 46-59 | 3711 (1367/2344) | 3421 (1274/2147) | 290 (93/197) | 7.8 (6.8/8.4) |
| 60-69 | 20198 (8535/11663) | 18387 (7847/10540) | 1811 (688/1123) | 9.0 (8.1 / 9.6) |
| 70-79 | 21440 (9621/11819) | 19351 (8801/10550) | 2089 (820/1269) | 9.7 (8.5/10.7) |
| 80-89 | 5095 (2268/2827) | 4469 (2012/2457) | 626 (256/370) | 12.3 (11.3/13.1) |
| > 89 | 409 (180/229) | 346 (152/194) | 63 (28/35) | 15.4 (15.6/15.3) |
| Total | 51187(22159/29028) | 46287 (20262/26025) | 4900 (1897/3003) | 9.6 (8.6/10.4) |

## Discussion

Currently, there is no method available globally that can be used for large-scale screening and can identify high-risk individuals for Alzheimer’s disease before clinical symptoms appear. Most blood testing methods use Aβ-PET as the gold standard, emphasizing consistency with Aβ-PET and aiming to become a surrogate tool for Aβ-PET. However, before Aβ-PET shows a positive result, Aβ has accumulated for 17-23 years ^[24]^, and by the time Aβ-PET is positive, regardless of whether there are symptoms of cognitive decline, the patient has already entered the preclinical AD stage ^[25]^. Therefore, current blood testing methods can be used for early diagnosis, but are difficult to use as early screening tools. The detection of urinary AβPP protein fragments is a real-time reflection of the Aβ content in the body, independent of Aβ-PET ^[20]^. Therefore, it has the potential to identify individuals with persistently elevated Aβ levels, i.e., high-risk individuals for Alzheimer’s disease, before Aβ accumulates to a positive PET result. In fact, previous reports have shown that the urine test strip can achieve a positive rate of nearly 80% in Aβ-PET negative MCI patients ^[20]^. In a community elderly population with 4,418 participants, the positive rate of the test strip increased with age, from 9.5% in people under 65 years old to 16.9% in people over 85 years old, which fully demonstrates its potential as an early screening tool ^[20]^. This characteristic of the positive rate increasing with age was further verified in this study: in the community population of 51,187 people, the positive rate increased from 6.3% in people under 46 years old to 15.4% in people over 89 years old. We also found that the positive rate of the test strip is closely related to cognitive ability. The scores of the two cognitive tests, MMSE and MoCA, were negatively correlated with the positive rate of the test strip. The lower the score, the higher the positive rate of the test strip, indicating a more severe degree of cognitive decline. Based on the above correlation between age and cognition, the test strip shows high feasibility and high accuracy as a screening tool for cognitive decline in a large population.

As a clinical auxiliary diagnostic tool, the positive rate of the test strips differed significantly between MCI/AD patients and healthy controls, with a κ value of 0.669, indicating good clinical concordance and early diagnostic value. This is very close to the previous report ^[20]^, showing its stable clinical performance.

Elderly people often have one or more sub-health conditions or comorbidities. These comorbidities, which are mainly in the periphery, may share some of the pathological change pathways as AD, or directly or indirectly affect the pathogenesis and course of AD ^[26]^. At the same time, treatment for these comorbidities may also improve the course of AD to some extent. To verify this idea, we used urine β-amyloid test strips as an assessment tool for the degree of cognitive decline and statistically analyzed the possibility of a single comorbidity leading to cognitive decline or Alzheimer’s disease. The results showed that memory decline was the most dangerous symptom, followed by hyperlipidemia and a history of stroke. This is consistent with the general consensus in the academic community ^[27]^. Our results showed that hyperglycemia did not significantly lead to cognitive decline, which may be due to the small sample size, or more likely because hyperglycemia is only one of the manifestations of AD, not the cause of AD. The fundamental pathogenesis of AD is insufficient mitochondrial energy metabolism, which leads to the inability to clear cellular waste in time ^[28]^. Another consequence of insufficient energy metabolism is the insufficient utilization of glucose, one of the energy storage substances, leading to hyperglycemia. Therefore, targeting blood glucose levels for AD treatment may have limited effectiveness. Drugs that effectively treat AD would necessarily improve mitochondrial function, thereby simultaneously treating hyperglycemia. From this perspective, test strips can also be used to assess the risk of AD comorbidities, undoubtedly providing a simple and practical tool for the pathological study of complex AD comorbidities.

Many elderly people need to take medication regularly to treat comorbidities. The medications could potentially have an improvement on cognitive decline. We analyzed the positive rates of test strips in patients with non-AD diseases and MCI/AD patients who had been taking a certain type of medication for a extended period of time. We found that patients with non-AD diseases who had been taking medication had a reduced positive rate of test strips to various degrees, and some were even lower than healthy people without any comorbidities. This verifies from one aspect that comorbidities and AD have a common pathological pathway. In contrast, the positive rate of test strips in patients taking AD treatment drugs did not decrease significantly, which also verifies the unsatisfactory performance of current AD treatment drugs. For example, the cholinesterase inhibitor donepezil hydrochloride was recently found to increase the risk of MCI patients’ disease exacerbation by 77% ^[29]^, which is consistent with our test strip results. If AD patients take anti-AD drugs while managing comorbidities such as hyperlipidemia, hyperglycemia and hypertension, the effect may be better than taking anti-AD drugs alone. In any case, these analysis results also provide a scientific basis for using test strips as a drug analysis tool with a unique perspective.

This study has several limitations. First, due to the insufficient clinical sample size and the complexity of comorbidities, we could only analyze the association of individual comorbidities, without performing multi-step, multivariate regression. Second, we only conducted a cross-sectional study and lacked long-term follow-up data, thus we could only assess the potential of the kit as an early screening tool for cognitive decline and Alzheimer’s disease. Our community-based mass screening in Yuhua District, Changsha, initially targeted 200,000 elderly people, but only slightly over 50,000 participated, and less than one-tenth of those who tested positive underwent follow-up testing. This indicates a lack of public awareness regarding the importance of AD, especially early intervention. There is a likelihood that some elderly people believe AD is a natural result of aging; have a fear of early screening; are unaware of the 20-30 year incubation period; or do not understand that diet and lifestyle can effectively delay the onset of AD. This underscores the urgent need to raise public awareness of AD, to ensure that the public fully accepts the concept of early screening and early intervention, and to better prevent and control AD.

## Data Availability

All data produced in the present study are available upon reasonable request to the authors

## Acknowledgments

We thank Qankorey Biotechnology (Changsha) for providing free test strips for the multi-center clinical trial. We also thank the People’s Government of Yuhua District, Changsha City, the Management Committee of Yuhua Economic Development Zone, the Health and Wellness Bureau of Yuhua District, and the Center for Disease Control and Prevention of Yuhua District for organizing free Alzheimer’s disease early screening and prevention work for people aged 50 and above in 14 community health service centers/hospitals within the district.

## Conflict of Interests

All authors declare no conflict of interests.

## Notes

### Competing Interest Statement

The authors have declared no competing interest.

### Funding Statement

This study did not receive any funding

### Author Declarations

Ethics Committee of Yuquan Hospital of Tsinghua University gave ethical approval for this work.

### Summary of Updates

Table 1, sepearated MCI+AD group to MCI group and AD group, so it is more clear to show the early screening value of the kit. Added Table 4-2, which shows the performance in cognitively normal participants only. This table further demonstrats the value of the kit in early stage of disease.

